# Active case finding using mobile vans equipped with artificial intelligence aided radiology tests and sputum collection for rapid diagnostic tests to reduce tuberculosis prevalence in rural China: protocol for a pragmatic trial

**DOI:** 10.1101/2024.12.08.24318678

**Authors:** Xiaolin Wei, Dabin Liang, Zhitong Zhang, Kevin Thorpe, Lingyun Zhou, Jinming Zhao, Huifang Qin, Xiaoyan Liang, Zhezhe Cui, Yan Huang, Liwen Huang, Mei Lin

**Affiliations:** Dalla Lana School of Public Health, University of Toronto, Toronto, ON, Canada; Guangxi Zhuang Autonomous Region Center for Disease Control and Prevention; Guangxi Key Laboratory of Major Infectious Disease Prevention and Control and Biosafety Emergency Response, Nanning, Guangxi Zhuang Autonomous Region, China

**Keywords:** Tuberculosis, active case finding, mobile van, artificial intelligence, cluster randomized controlled trial, China

## Abstract

**Background:** Tuberculosis (TB) remains a significant public health challenge, particularly in rural areas of high-burden countries like China. Active case finding (ACF) and timely treatment has been proved effective in reducing TB prevalence but it is still unknown regarding the impact on TB epidemic when employing new technologies in ACF. This study aims to evaluate the effectiveness of a comprehensive ACF package utilizing mobile vans equipped with artificial intelligence (AI)-aided radiology, and GeneXpert testing in reducing TB prevalence among high-risk populations in rural Guangxi, China.

**Methods:** A pragmatic cluster randomized controlled trial will be conducted in two counties of Guangxi, China. The trial will randomize 23 townships to intervention or control groups at 1:1 ratio. The intervention group will receive a single ACF campaign in Year 1, incorporating mobile vans, AI-based DR screening, symptom assessment, and sputum collection for GeneXpert testing. Control group participants will receive usual care. TB patients identified in Year 1 will be required to complete TB treatment in Year 2. The primary outcome is the prevalence rate of bacteriologically confirmed TB among high-risk populations in Year 3. Process evaluation will explore adaption, acceptability and feasibility of the intervention. We will conduct incremental costing study to inform future scale-up of the intervention in other settings.

**Discussion:** This study will provide valuable insights into the effectiveness and feasibility of utilizing AI, mobile vans and GeneXpert for TB ACF to reduce TB prevalence in rural settings. If successful, this model will contribute to possible solutions to achieve the WHO End TB Strategy by 2035.

**Trial registration**: ClinicalTrials.gov Identifier -NCT06702774

## Introduction

The United Nations passed the Resolution to end tuberculosis (TB) by 2035. TB is the leading cause of death in infectious disease. In 2022, the world reported 7.5 million new TB cases and 1.3 million deaths caused by TB [1]. TB incidence has declined by 2% a year between 2010 and 2020. However, the rate has reverted to increase by 3.9% between 2020 and 2022 due to the COVID-19 pandemic [1]. To reach the END-TB resolution, global TB incidence has to be reduced to less than 100 per million by 2035 [2]. The world is looking for evidence of innovative approaches to eliminate TB in a relatively short time frame [3].

The principle of “prevent, search, detect, treat” that has successfully reduced TB prevalence in high-income countries in the last century [4]. Studies in high TB burden countries also proved that active case finding (ACF) at the community level would identify patients who are in their early stage of disease development, or who may be delayed or missed in the routine passive case finding at health facilities [4, 5]. A 2009 study in rural Zimbabwe showed both mobile van and household-to-household sputum collection methods lowered TB prevalence by 41% in three years, showing the potential of using mobile vans in case finding [6]. Another study in rural Vietnam demonstrated that conducting annual community screening based on a rapid molecular diagnostic tool (GeneXpert) for three years could reduce 40% of TB prevalence in a high TB burden setting [7]. However, GeneXpert costs around US$15 per test, which is prohibitive to be used as a large-scale screening tool.

More cost-effective active case-finding tools, such as computer-aided diagnostic tools using artificial intelligence (AI) deep learning neural networks, have shown satisfying sensitivity (86%) as a screening tool to reduce 66% of cases needed for laboratory tests [8]. Operational studies revealed the feasibility and effectiveness of using AI-facilitated X-ray and GeneXpert for community screening and diagnosis [9]; however, most studies were pre-after comparisons that were not properly designed to evaluate epidemiological impacts [10]. In addition, two studies in Africa highlighted that the interventions were more cost-effective when screening elders, close contacts with active TB patients, and individuals with comorbidities such as diabetes and HIV [11, 12].

Here, we aim to implement and evaluate the feasibility and effectiveness of a comprehensive intervention package in rural Guangxi, China, a high TB prevalent setting, that employs the latest technologies for active case finding to reduce TB epidemics among high-risk populations. The technologies include employing a mobile van equipped with AI aided radiology screening and GeneXpert test for community active case finding in communities. Our study is designed as a pragmatic clustered randomized controlled trial to evaluate the effectiveness of the comprehensive package in reducing TB prevalence over a period of three years among high-risk populations.

## Methods

This protocol is reported according to the Standard Protocol Items: Recommendations for Interventional Trials (SPIRIT) guidelines (see S1 Table).

### Study status and timeline

The lifespan of this study will be 42 months in total, including 36 months for the main trial (Year 1-3). More details of trial timeline are included in the Fig 1. The trial has been started since Nov 2021 and has been suspended for 3 months due to COVID-19 control in Year 1, thus the progress was postponed accordingly. Participant recruitment and data collection has been ongoing by the time of submission of this protocol, which are estimated to be completed by end of January 2025. The trial results will be disseminated through research articles and policy briefs after the trial evaluation is completed after April 2025. We aim to publish our results and findings in leading international medical journals and present at national and international conferences.

**Fig 1.**
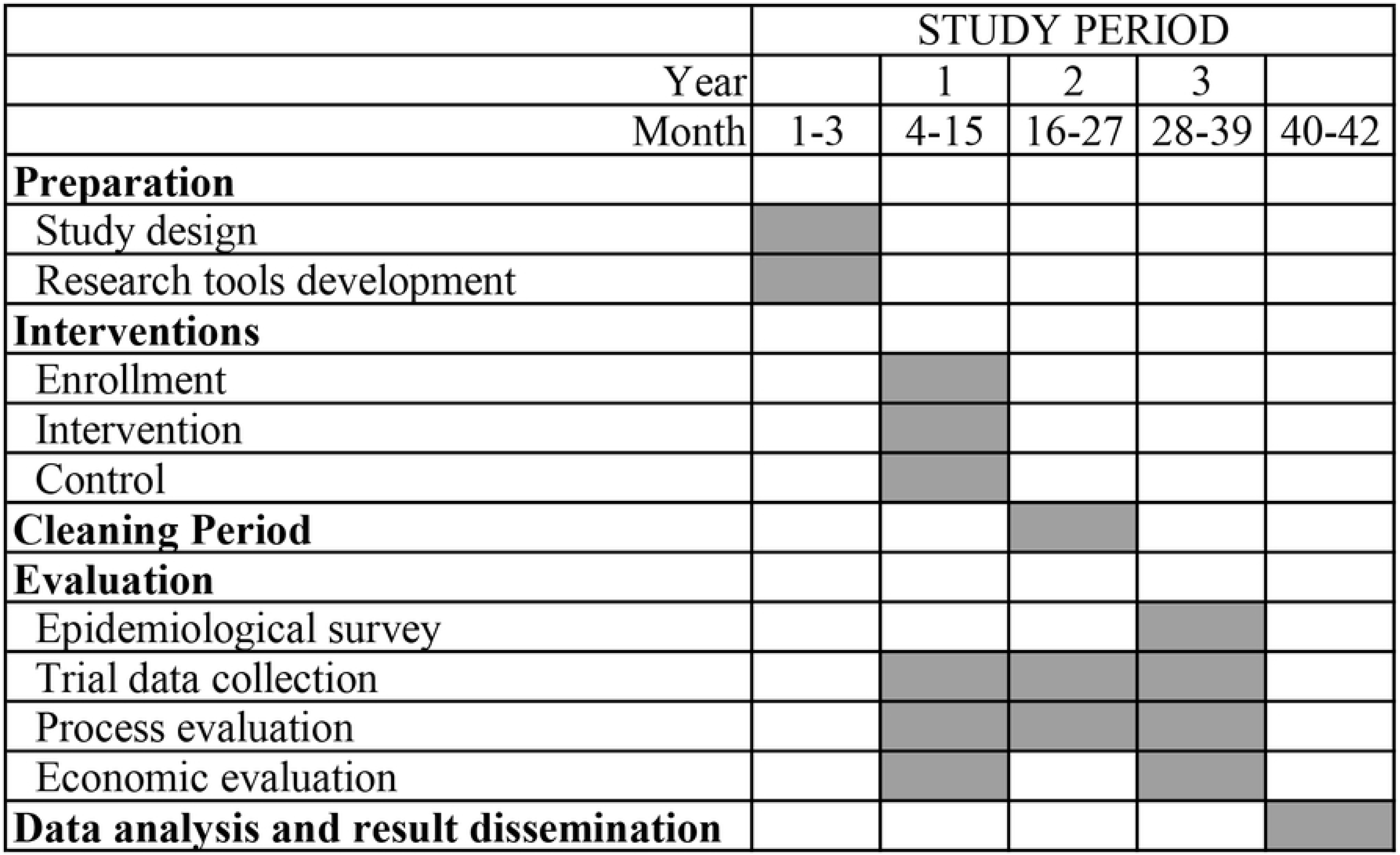
SPIRIT schedule of enrolment.

### Study design and setting

We will conduct a parallel two-group, cluster randomized controlled trial of a comprehensive package of active case finding followed by timely treatment in rural Guangxi China to evaluate whether the interventions could reduce the TB epidemic compared to the usual care (Fig 2). Guangxi is located in southwest China bordering with Vietnam. Guangxi reported 31,913 TB cases with notification rate of 70/100,000 in 2020, representing the second highest TB burden in China. Of the reported TB cases, 74% were from rural areas, and 30% were elderly (≥ 65 years old). Mountainous areas in Guangxi had high TB notification rates; for example, the two trial counties (Xincheng and Xiangzhou of Laibin Prefecture) reported an average notification rate of 134/100,000, which is two times the provincial average. In addition, the Covid-19 pandemic has substantially delayed TB diagnosis and treatment, which would fuel up TB transmission and exacerbate TB epidemic.

**Fig 2.**
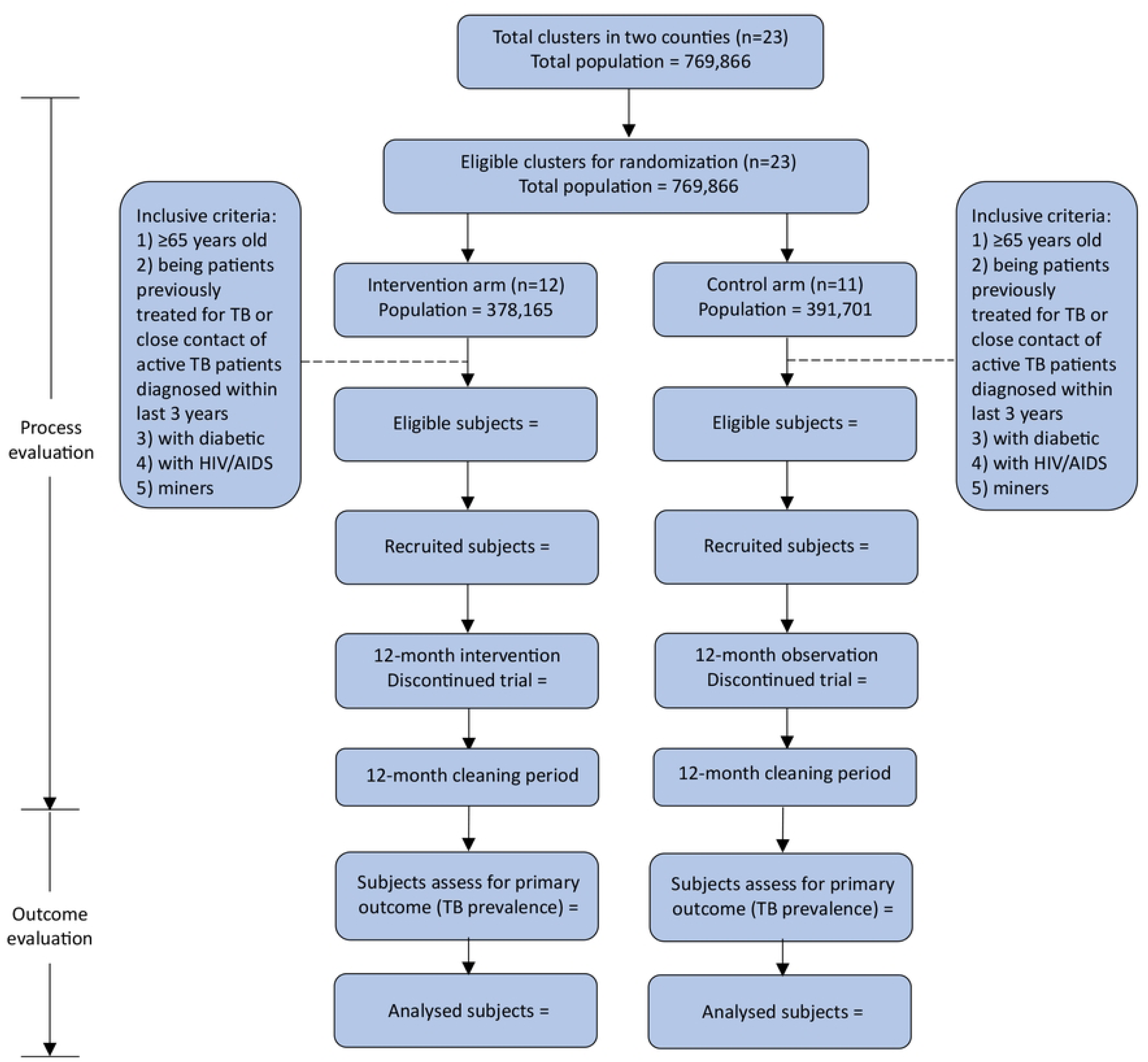
Trial design flow chart.

TB services in China is funded and delivered by the public health system at the county, township and village levels. The TB free treatment policy requires basic TB diagnosis tests and treatment should be free (for drug susceptible cases) or supported by the health insurance schemes (for drug resistant cases). The County Centre for Disease Control (CDC) is responsible for TB planning, reporting and quality control, while TB clinical care is provided by the TB Clinic at the County Hospital. Passive case finding is the practice of usual care, where patients with chronic cough or other TB-related symptoms are self-presented or referred to the County Hospital for subsequent diagnosis and treatment. TB diagnosis is based on sputum smear, chest X-ray examinations, and clinical assessments. Standard WHO DOTS treatment regimen is provided for patients diagnosed of drug-susceptible TB. TB patients visit the County Hospital monthly for medication renewals. Patients take their anti-TB medications on a daily basis with the support of village doctors, who are community health workers. In Guangxi, village doctors are semi-private who receive 50% of income from the township hospitals for their public health work such as maternal and child health, immunization, pap smear screening, and disease management including TB, diabetes, and hypertension, while they earn the other 50% income from clinical consultations under the rural health insurance scheme. According to the National TB Guideline, village doctors are responsible to visit TB patients weekly. The public health doctor in the township hospital is required to supervise village doctors and visit patients monthly. However, the actual frequency is far less and often substituted via phone calls. In each village, there is a female social worker in the Village Committee, who plays a major role in public education, maternal and child care, and protecting women’s rights. These female social workers have a strong connection with villagers but have not been involved in TB care.

### Eligibility criteria

#### Clusters

We have two levels of clustering in this trial. Township is treated as the cluster of intervention. A township is an administrative unit with a township hospital and consists of 7-12 villages. All townships in Xincheng and Xiangzhou county, which provide their willingness to participate, will be included. Villages within the participating township serve as sub-cluster in the survey that determines the TB prevalence rate.

#### Participants

In each township, all residents who are 15 or older become eligible and defined as high-risk populations if they: 1) are elderly (i.e., aged 65 and above), or 2) been aged below 65 but with one of the following conditions: being patients previously treated for TB or close contacts of a patient with a TB patient diagnosed within the last three years; having been clinically diagnosed with diabetes, HIV positive, or worked as a miner. All eligible participants who signed informed consent form will be recruited to the study. Residents who refuse participation will be excluded.

### Procedures

#### TB Care provided in both groups

Routine TB care is provided per the China National TB Guideline to both groups, where passive case finding is practiced. Patients self-present or are referred to the TB clinic at the County Hospital when they seek care for TB related symptoms. TB diagnosis is based on X-ray examination, sputum smears and clinical assessment. Confirmed TB patients are treated in County Hospitals based on the WHO DOTS regiments for drug-sensitive TB. Those who are diagnosed with multi-drug resistant (MDR) TB are referred to prefecture TB designated hospital.

#### Usual care

Usual care will be provided in the control group, and no active case finding activities will be provided.

#### Intervention

In the intervention group, in addition to usual care, we will implement one active case-finding campaign among all eligible participants in Year 1 (Fig 3). Villagers will be informed of the screening through public information and social workers. Prior to the visit, village social workers and village doctors will conduct door-to-door visits to recruit participants and obtain their informed consent. We will employ a mobile van equipped with the AI-facilitated digital radiography (DR) machine and refrigerator to visit villages on a mutually agreed date. On the day, all eligible participants will be invited to complete a brief questionnaire regarding TB related symptoms, and then invited to have their DR examined in the van. Those who are presumptive of TB based on either DR examination or any TB related symptoms, including any chronic cough over two weeks, hemoptysis, fever, night sweats, chest pain or unexplained weight loss, will be asked to produce a single on-site sputum. These villagers are also asked to take two specimen boxes for collection of night and morning sputum. All staff are trained for good quality sputum sample collection, storage and transportation. Medical nebulizer inhalation will be provided to those who can not provide good quality sputum. Sputum samples, either collected on the day or delivered by village doctors within 48 hours, will be stored in the refrigerator of the mobile van and transported to the county hospitals at daily basis for sputum smear, culture and GeneXpert using sample mixed approach which method of mixing is published elsewhere [13]. Participants who are bacteriologically negative but have abnormal DR result or TB symptoms will be referred to the county hospital for clinical assessment based on China National TB guideline. All diagnosed TB patients, either bacteriologically or clinically confirmed, will be notified by the village doctor within 48 hours and referred to the County Hospital for standard WHO DOTS regiments per national guideline.

**Fig 3.**
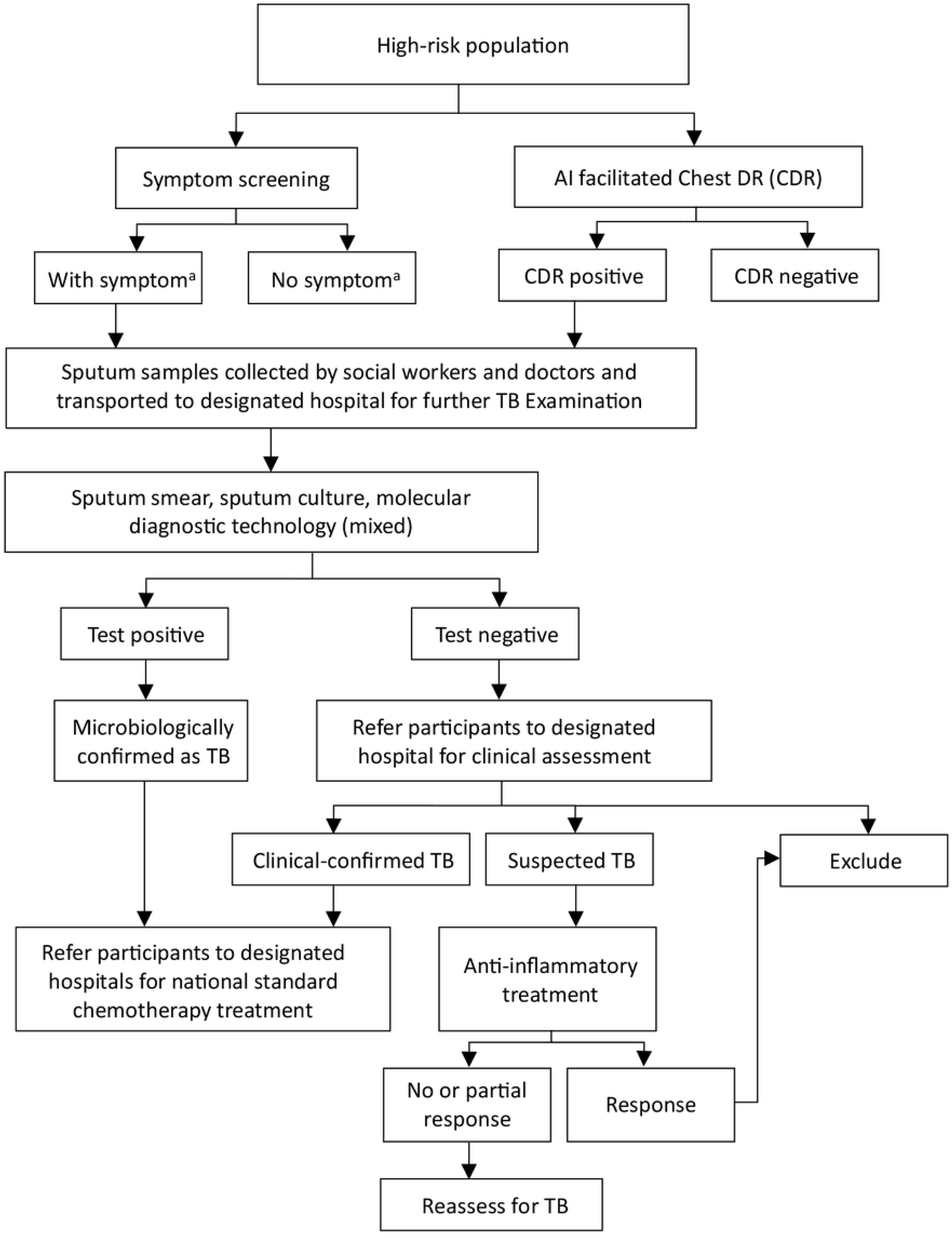
Algorithm of active TB case finding in intervention group. a. TB symptoms include chronic cough over 2 weeks, hemoptysis, fever, night sweats, chest pain or unexplained weight loss. TB= tuberculosis, DR= digital radiography

#### Research activities apply to both groups

We will wait for a year (cleaning period) to ensure all TB patients identified in Year 1 complete the treatment. We will conduct epidemiological prevalence survey in both groups in Year 3 using cluster-random sampling methods adopted from WHO [14], targeting high-risk populations in both intervention and control groups, to compare the TB prevalence rates. Regarding the survey, we will apply the same active case finding activities as outlined above to both intervention and control groups. For feasibility reasons, we plan to conduct the survey in Year 3 at the village (sub-cluster) levels in both groups.

### Outcomes

Our primary outcome indicator is the prevalence rate of bacteriologically positive TB in Year 3 among the high-risk populations, including those 65 and older, those who are under 65 but have a history of tuberculosis treatment or have been in close contact with a person diagnosed with TB within the past three years, have been clinically diagnosed with diabetes, HIV, or have a background of working as a miner. According to WHO definitions, bacteriologically positive case is defined as having a positive test from any of the three tests (i.e., GeneXpert, smear or culture). Prevalence is estimated using a cluster-random survey among the high-risk populations that is conducted in both the intervention and control groups in Year 3.

There is a list of secondary outcome indicators, which allow us to assess and evaluate whether our interventions could bring positive impacts on TB epidemic in the intervention and control groups: 1) prevalence rate of active TB, including both bacteriologically positive and negative cases, among the high-risk populations in Year 3. This indicator will also rely on estimates from the epidemiological survey in Year 3; 2) notification rates of bacteriologically positive TB cases among all populations in Year 3; and 3) notification rates of active TB cases, including both bacteriologically positive and negative cases among all populations in Year 3. Nominators of the two notification rates will include TB cases identified from our epidemiological survey in Year 3 plus TB cases reported from the National TB Program, while both denominators include all adult residents (i.e., those of 15 years and older) in the intervention and control groups.

We will also collect costs from patient, health system and societal perspectives to conduct an incremental costing study. We will collect process indicators that will including a) the proportion of bacteriologically positive cases identified by active case finding in the intervention group; b) the yield rate of our intervention, which is the number of bacteriologically positive cases per 100 persons screened using each strategy.

### Sample size

The sample size is calculated for the investigation of the primary outcome, i.e, the prevalence rate of bacteriologically confirmed TB, that will be conducted in Year 3. We estimated that the TB prevalence rate among the high-risk populations in the trial area is about 300 per 100,000. We assume our intervention will reduce the rate among the high-risk populations in the intervention group by 50% (i.e., to 150 per 100,000) within 24 months, which can be considered as a significant reduction at the population level. We estimate that the intra-class correlation coefficient for TB prevalence was 0.001 [7]. The average number of eligible participants is about 500 per village based on the initial reports. We estimate that we would need 47 villages (that is, approximately 23,500 participants) in each group to have 80% power to detect the reduction estimated above, assuming p < 0.05.

### Recruitment, randomization and blinding

There are 23 townships and 255 villages in Xincheng and Xiangzhou county of Laibin city [15]. We will implement the active case finding intervention at the township level to avoid contamination because township hospitals manage village doctors. We seek to recruit all willing townships. After recruitment, the study statistician will use a custom-written computer program to randomize all recruited townships, stratified by county, assigning them to the control and intervention group in an overall 1:1 ratio (12 intervention vs 11 control) [16]. Then township/village doctors and county hospital staff in the intervention group will be invited to receive training about specific interventions in this project. For the epidemiological survey in Year 3, the statistician will randomly select 47 villages (sub-clusters) from each group stratified by county. We will seek written informed consent from each participant before enrollment, as we need to collect their health information, bio-sample (sputum) and test result for research purpose.

Due to the nature of the intervention, neither single nor double blindness procedure is applicable. However, we will blind the data analysts who process the end-point data for outcome assessment.

### Data collection

We will collect all types of field data produced during the trial, including patients’ basic information, disease history, health information, DR test result, laboratory test result for TB and diagnosis, as well as routine TB reporting data. All data will be entered into a data website designed for this project, accessible only by designated researchers via local intranet with a password. Individual data will be deidentified and stored for five years after the completion of the trial. For research purpose, we will define all outcomes at the subcluster-level and will calculate them from the individual-level data as single summary outcome values, such as prevalence per cluster.

#### Active screening

During the active case screening, we will collect the information about suspicious symptoms, including chronic cough for more than 2 weeks, hemoptysis, blood sputum, fever, night sweats, chest pain or unexplained weight loss, and TB risk factors (close contact history of tuberculosis patients, diabetes, AIDS/HIV and dust exposure).

#### Laboratory testing and clinical consultation

The county hospitals will carry out laboratory testing according to the ‘TB Laboratory Testing Protocol’. Sputum smear test, GeneXpert test and sputum culture are carried out at the same time. Every five samples of sputum are mixed for GeneXpert test to save costs. Those positive mixed sample will be tested separately to confirm the positive case. Country hospitals will distribute the laboratory testing results to patients within 48 hours. All TB diagnoses will be recorded in the ‘Register of Newly Diagnosed Patients’ (labelled with subject number) in detail. Clinical diagnosis based on the clinical assessment is performed by doctors in county hospitals according to the national TB Classification Criteria (WS196-2017) and Diagnostic Criteria (WS288-2017) [17, 18]. We will also collect TB cases diagnosed through the usual care in both intervention and control groups through the three years.

#### Primary and secondary outcomes

The data collection approach in Year 3 for the outcome evaluation will be the same as the active TB case finding in the intervention group in Year 1. As we define most of our outcomes as single summary values at the subcluster-level, such as prevalence and incidence, we will sample from both intervention and control group, calculate and compare the outcomes following the trial period. Take the primary outcome, the prevalence of bacteriologically positive TB, for instance, we will sample all high-risk populations of selected villages (sub-clusters) of the two groups, conduct a sputum smear, GeneXpert, sputum culture test, and compare the calculated prevalence of bacteriologically positive TB. A similar process also applies to measure secondary outcomes, such as prevalence of active TB in two groups.

### Statistical analysis

The results of this project will be reported according to the ‘Consolidated Standards of Reporting Trials: Extension for Cluster Trials’ (CONSORT) guidelines [19]. We will create a detailed statistical analysis plan. In the full research paper, we will present appropriate descriptive statistics for the two counties and all relevant participants before and during the trial. In addition, using cluster-level methods of analysis, we can get a main set of estimates for our primary and secondary outcomes after the trial, acting as the primary evidence for determining the effectiveness of this trial. Appropriate summary statistics and their associated 95% confidence interval for all differences outcomes between the intervention group and the control group will be calculated. No interim analyses are involved in this project.

For the primary outcome and all secondary proportion-based outcomes, the statistical analysis will employ a mixed-effect logistic regression model including random effects for township and village. The intervention effect will be expressed as an odds ratio with 95% confidence intervals. Secondary analyses of these outcomes will adjust for the following variables: age, sex, and ethnicity.

For continuous secondary outcomes we will employ a linear mixed-effect model with random effects for township and village. This model will also adjust for the following variables: age, sex, and ethnicity. The intervention effect will be expressed as the adjusted mean difference with 95% confidence intervals.

### Process evaluation

Since our project contains multiple components, the process evaluation during and after the trial is guided by the framework for design and evaluation of complex interventions [20, 21]. Utilizing a combination of qualitative and quantitative methods, process evaluation for the complex interventions in our cRCT can explore the mechanism of achieving the intervention effects [22]. In other words, we can get insights of a series questions such as ‘what interventions are effective in which policies and implementation environments, for whom and why?’. We will conduct around 30 in-depth semi-structured interviews with leading personnel at all levels of health agencies, township and village doctors, county hospital staffs, social workers, and trial participants in the intervention group.

Our specific objectives for the process evaluation include: 1) to determine intervention accessibility by the number of subjects receiving screening, presented to county hospitals, and under treatment, as well as their feedbacks; 2) to explore the intervention effectiveness (reasons for both effectiveness and ineffectiveness), along with the relationship between interventions and expected results; 3) to access the acceptance of intervention implementation at the organization level and the staff level – any conflicts with existing management or policies, and corresponding adjustment; 4) to report intervention fidelity, the degree and quality of intervention completion, as well as challenges and unexpected results, at both subcluster-level and individual level.

### Analysis of process evaluation

All the recorded interviews will be transcribed and translated into English before qualitative analysis. We will use NVivo 10 software to process and analyze our qualitative data guided by a framework analysis as described by Gale at al [23], to identify themes related to our process evaluation objectives. The findings will be reported based on the Consolidated Criteria for Reporting Qualitative Research (COREQ) guidelines [24].

### Costing study

This study will measure the incremental cost-effectiveness of interventions compared with conventional TB finding, using research data and literature to evaluate both health system and social benefits. We will compare direct costs and outcomes of the intervention group to those of the control group throughout the 24-month trial. The objective of the costing study is to calculate the incremental cost-effectiveness ratio (the increased costs required for each 1% reduction in the prevalence of active TB among high-risk populations) using data from the project’s primary outcome.

#### Data collection

We will send out a questionnaire to directors of all recruited township or village hospitals to collect health care resources used in the trial, along with their costs.

The data include: 1) average salaries, in RMB, for each level of staff in designed hospitals who participated in this project; 2) average salaries, in RMB, for township and village doctors who do not work in designated hospitals but still participated in the project; 3) average salaries, in RMB, for social workers and other personnel worked in the project implementation; 4) the duration of consultations on mobile health vehicle and in county hospital); 5) the frequency and duration of staff training and sequential meetings during the process; 7) cost for TB screening (i.e. symptom screening, chest DR, mobile van operation, sending samples to county hospitals) and laboratory test (e.g. sputum smear, culture, and GeneXpert). To optimize the data quality, we will double-entry and random-check the input data.

#### Estimation of resources use and costs

Healthcare resources in this case include resources used for TB detection and diagnosis. The cost per patient is calculated as the sum of consultation, screening and laboratory testing. The total costs is calculated for the main analysis population as described in the Statistical Analysis section [25].

#### Estimation of implementation costs

The implementation costs are calculated separately and will not be accounted in the cost-effectiveness analysis. The objective of this estimation is to inform policy makers and stakeholder of the potential cost to facilitate their decision on whether to implement the intervention at scale. For each cluster, the implementation costs included the cost of training for personnel, the cost of personnel to attend training, the costs for developing and printing the handbook or guidelines per training attendee, operation cost of mobile van, driver cost, setup cost of AI facilitated DR, the costs for producing symptom questionnaires used in the project, and the costs of sending samples back to county hospitals.

### Trial management

Guangxi CDC will lead this project and have full access to all the anonymized data and information associated with the trial. We will establish a data monitoring committee (DMC) to ensure all data are collected in conformity with the agreed ethical guidelines, stored in Guangxi CDC properly and used for research purposes only. Meanwhile, the DMC is also responsible for the safety and privacy protection of all the subjects and providers participated in the trial. Additionally, there will be a trial steering committee (TSC) lead by external members to generate the progress of the trial. We will hold regular online meetings with both DMC and TSC every 6 months to get updates and discuss any protocol modifications, starting from the beginning of the study, until it completes. If there are any changes or concerns needed to report or discuss, the committees can meet on an ad hoc basis with the co-leaders as well. Moreover, a trial management unit (TMU) will be established in consisting of three local staff. The responsibility of the TMU is to better manage the day-to-day activities of the trial according to the study protocol.

## Discussion

This study aims to enhance the existing TB control protocol in an area of high TB prevalence. As the location of implementation, Laibin is an ethnically diverse area with the highest reported TB incidence rate in Guangxi region. In other words, it has an urgent need to find a more effective way to manage its TB epidemic. Given that about 27% of all reposted cases in Guangxi region are from the elderly population, the interventions target the elderly and also pay special attention to other populations who have high risk of TB infection [15, 26, 27].

The study has significant policy relevance to China and globally. The objective of this study is to effectively reduce the TB epidemic in Guangxi and achieve the WHO’s End TB Strategy [2] milestones by 2024. If successful, the prevalence of TB in the intervention areas will be reduced by more than 50%. It aligns with the goal to build a healthy China as well. The successful implementation of this study will yield a feasible TB control strategy that can be replicated in other parts of China, which may provide a model for countries with similar context to achieve to the WHO’s goal to end TB by 2035 [2].

Furthermore, active screening interventions involve innovative technologies. Since over 40% of areas in Laibin are mountainous, the mobile health vehicles equipped with DR machines and refrigerators make active screening procedures more convenient, and all the bio-samples can be properly stored and timely transported to the county hospital. In addition, the AI-based diagnostic system (above 90% sensitivity) offers axillary support to detect abnormalities in DR images, increasing the test efficacy [28].

Nevertheless, there are some limitations in this study as well. First, TB-suspected participants in the active screening intervention are required to collect two sputum samples (one for morning and one for night sputum) and send them to village doctors. Although specific equipment and instructions will be given, there are still risks of improper sample collection or transportation, which may affect laboratory results and TB diagnosis. We provide training for people assumptive of TB in sputum collection to improve the quality. Secondly, due to the limitation of funding and human resources, we are not able to conduct active case finding among all populations in participating townships in Year 3. Therefore, the trial cannot evaluate the impact of interventions on TB prevalence in the general population. On the other side, a survey of the population may not be feasible anyway because most people of working ages in rural areas moved to urban areas as migrants during most time of the year, while only elders, who comprise the majority of our high-risk population, and children stay at home. Our active case-finding activity targeting high-risk populations would significantly reduce TB in rural areas.

## List of abbreviations

AI: Artificial Intelligence
AIDS: Acquired Immunodeficiency Syndrome
CDC: Center for Disease Prevention and Control
DMC: Data Monitoring Committee
DR: Digital Radiography
HIV: Human immunodeficiency Virus
MDR-TB: Multidrug-resistant Tuberculosis
NTP: National TB Program
RCT/cRCT: Randomised Controlled Trial/Cluster Randomised Controlled Trial
TB: Tuberculosis
TCS: Trial Steering Committee
TMU: Trial Management Unit
WHO: World Health Organization

## Acknowledgement

We acknowledge Guangxi Laibin Prefectural Center for Disease Control and Prevention, Xincheng and Xiangzhou County Centers for Disease Control and Prevention as well as all the health providers and patients who participate in discussions to develop the proposal.

## Data sharing statement

Metadata won’t be publicly available due to local policy. Clustered data can be shared by email (Guangxi CDC) based on reasonable request for research purpose only upon approval after January 1, 2027.

## Ethical approvals and consent to participate

The study has obtained ethical approval from the Guangxi Institutional Review Board of the Guangxi CDC on 26 August 2021 (Ref: GXIRB2021-0033). Written informed consent form will be collected before any patient is recruited into the study.

## Supporting information

S1 Table SPIRIT checklist

